# On the true numbers of COVID-19 infections: behind the available data

**DOI:** 10.1101/2020.05.26.20114074

**Authors:** M.E. Qadmiry, E.H. Tahri, Y. Hassouni

**Affiliations:** Laboratory of Matter and Radiations Physics, Faculty of Sciences, University of Mohamed I, Av. Mohamed VI, BP 717, Oujda, Morocco; ESMaR, Faculty of Sciences, Mohammed V University, Av. Ibn Battouta, B.P. 1014, Agdal, Rabat, Morocco

## Abstract

In December-2019 China reported several cases of a novel coronavirus later called COVID-19. In this work, we will use a probabilistic method for approximating the true daily numbers of infected. Based on two distribution functions to describe the spontaneous recovered cases on the one hand and the detected cases on the other hand. The impact of the underlying variables of these functions is discussed. The detected rate is predicted to be between 5.3% and 12%, which means that there would be about 68 million infected until now (25-May 2020), rather than the officially declared number of 5.37 million worldwide cases.

## 1 Introduction

Since the outbreak of the novel coronavirus COVID-19 and its spread around the world, several published works were concerned with the analysis of the available data to extract information that characterizes the daily evolution of this new pandemic. Among the concerned information, there are the incubation period[1,2], the reproduction number[3], mortality rate[4], and the asymptomatic proportion as in [5], where was used the data of the Diamond Princess cruise ship; which means that the study is done on a closed population, a fact that is worthy to mention.

Our aim in this work is to approach the true number of infected cases, and to develop an analytical method in order to allow the simulation of different scenarios that can occur if we modify the underlying variables of two special probabilistic functions. These probabilistic functions control the numbers of the infected as well as the numbers of the detected cases. To predict the evolution of the true number of cases; which escapes from the official counting, we take into account the reproduction number.

The importance of this investigation lies in the fact that the important part of the infection comes from people who are not in hospitals or not isolated, i.e., that the virus transmission occurs in general before symptoms appear [6].

Then, we think that the numerical simulation achieved here can be helpful to estimate the consequences of different strategies adopted at the governmental level, such as social distancing, strict-confinement for a short period, mass testing, and other measures that aim to stop the evolution of the pandemic.

## 2 Method

In this section, we will introduce the distribution functions that describe the spontaneous outcome cases and cases detected. In addition to the reproduction number responsible for exponential growth.

### 2.1 Spontaneous outcome cases

When there is no medical intervention in favor of the infected, the biological defending system has to withstand alone as a natural solution. Therefore, eventually the cases end by either recovering disability or death. Thus, for a closed population initially infected, the number of infected keeps steadily decreasing since the illness onset until the end of infection, concerning many factors including age, health status, viral load, immunity, etc.

Considering these facts, we will suggest a suitable distribution function to lower the amount of the infected population through time. It is denoted as *Ps(t)* -(s)-subscript as survival- and its analytical expression is chosen so as to be compatible with the empirical observation taken from the data available in [7] based on 1987 control patients, so it is given as follows:

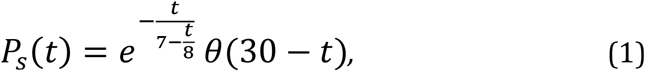

where *θ*(*t*) is the Heaviside step function equals to 1 if *t* ≤ 30 and to 0 otherwise, it ensures here that the function vanishes if the time exceeds the period of 30-days since the illness onset. This is because there is no remaining infected after this period, which after all, we will see that it does not have a significant impact on the results. Let us note also that we do not use the polynomial regression to describe *P_s_(t*), because it demands a greet polynomial degree.

In Fig: (1), we show the behavior of the *P_s_(t*) function. The real curve can be extracted from the data concerning the control patients under the drug treatments experiment as for example in [7,8], here we use the data of the first one as we mention above. The comparison is shown in Fig: (2), where we take the initial number of infected to be 1987. Thus, we plot the expected survival cases -at every day-given by the product 1987 × *P_s_(t*), with the real survival cases following the data. The standard error equals to 19, which means that we have a good approximation.

**Figure 1:**
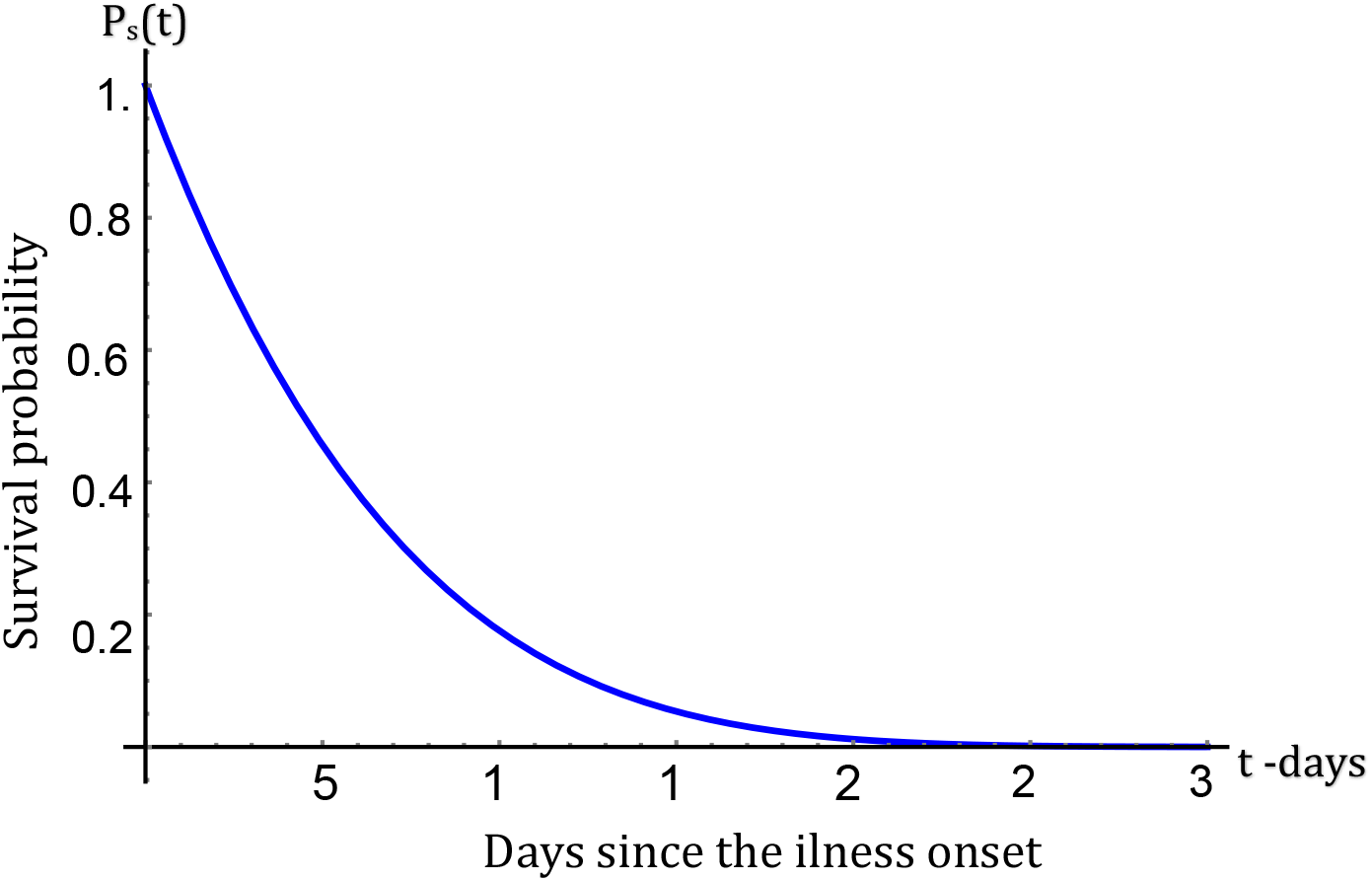
The behavior of *P_s_(t*) function

**Figure 2:**
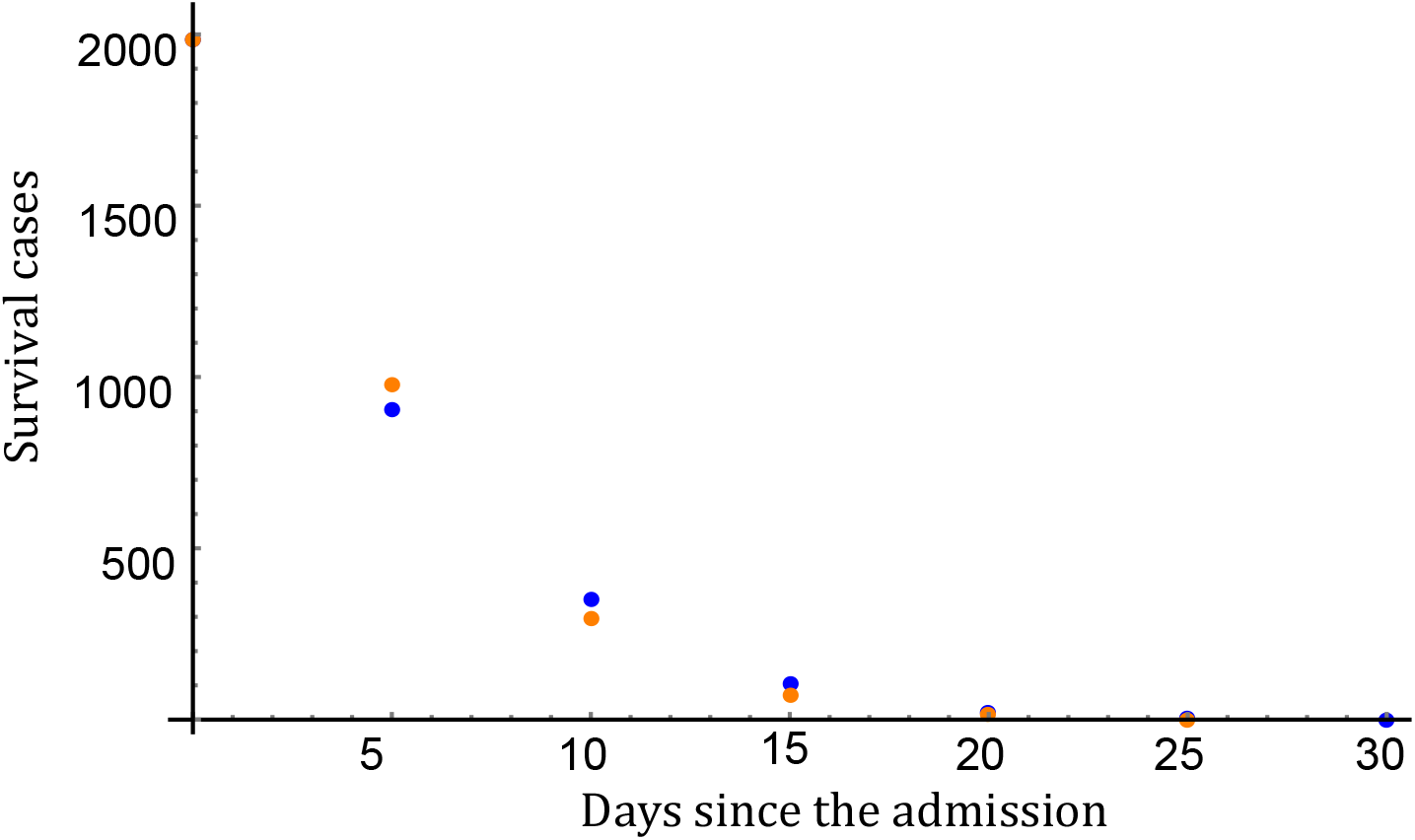
Real (orange) and expected (blue) survival cases using the available data and the *P_s_*(*t*) function, respectively

### 2.2 Detected cases

There is no doubt that the cases counting is not perfect, because there are many factors that may affect the results, such as test accuracy; viral load; medical protocol; testing strategy… Nevertheless, we can estimate another cumulative distribution function (CDF), which describes the probability to detect the infected cases among the entire population. This function is well-known used for several precedent epidemics (see [9]), e.g., SARS coronavirus; Rhinovirus; Influenza B…

The logistic distribution is useful to recognize this CDF, it is characterized by two parameters: the first one, denoted by *M*, represents the maximum absolute accuracy of testing and the second one is denoted by *δ*_0_ and represents the incubation period of COVID-19, which is estimated to be between 5.5 and 9.5 days (see [5]), e.g. it has fixed to 5.8 days in [10] and 5.1 days in [2]. Let us denote this distribution function by *P_d_(t)* -(d)-subscript as detected, and "t” as time representing days since infection- and write its explicit expression in the following manner:

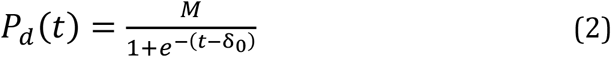

In what follows, we will choose the value of the incubation period as *δ*_0_ = 6.4-days following the result of the reference [11] (where it was chosen for the accuracy of the data used in the analysis). Concerning the testing accuracy, we take the maximum equal to *M* = 90%, which is a widely varied quantity regarding the testing strategy and the asymptomatic proportion estimated in [5] to be 17.9%.

In Fig:(3) we show the evolution of *P_d_(t)* CDF with respect to the above conditions, from which we get the probability to detect 2.9% after 2 days; 58% after 7 days; 90% after 12 days.

**Figure 3:**
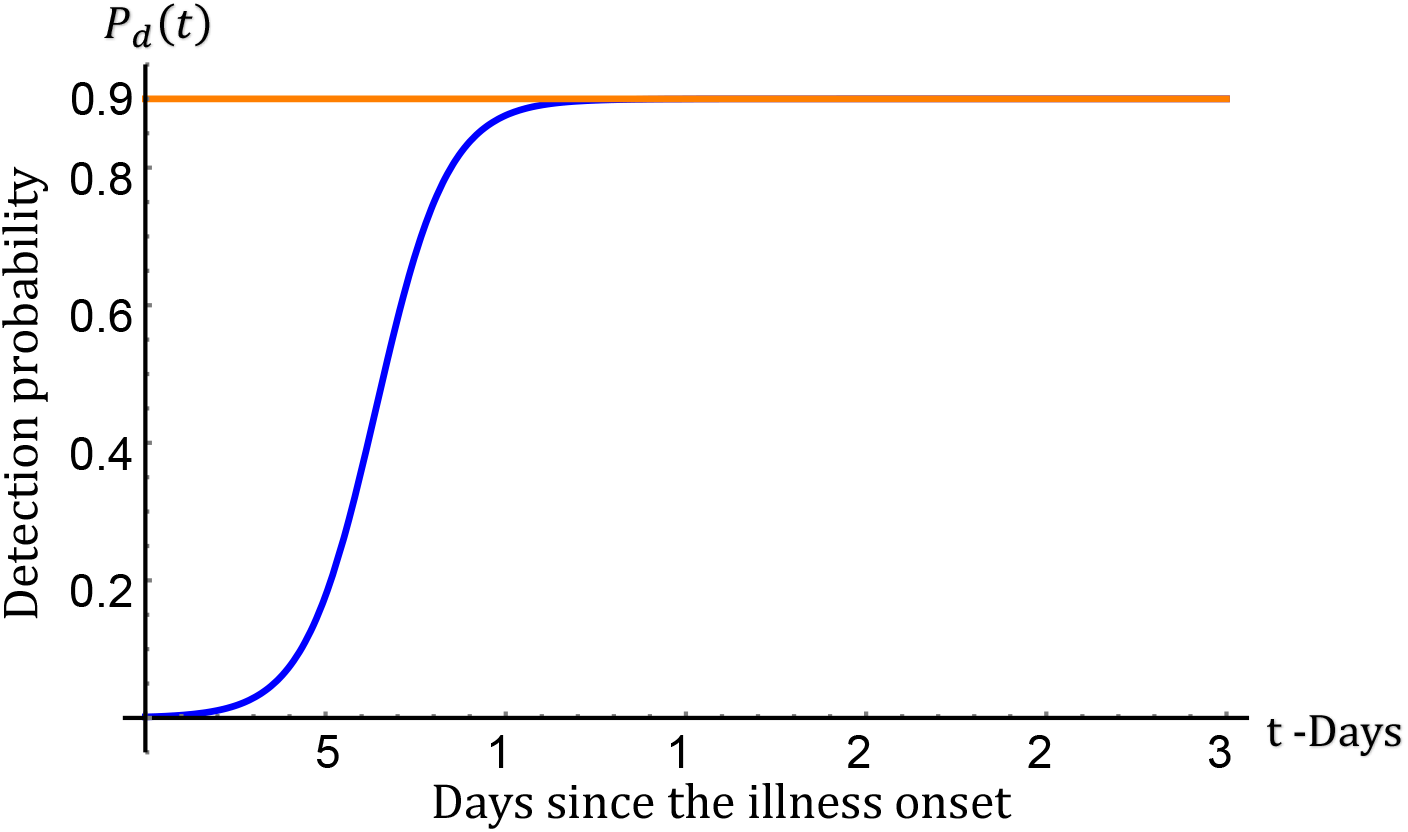
Graphic illustrating the behavior of the *P_d_(t)* CDF by considering M=90% and *δ*_0_ = 6.4 days

### 2.3 Evolution of reproduction number

The reproduction number is an important index to evaluate the spread of an epidemic. It is defined as the number of infected caused by an infectious person during the infectious period, denoted by *R*_0_.

What interests us in this work is the infection-producing contacts per one day, which is described by a function of time denoted by *r(t*), obtained simply as the ratio of the reproduction number *R*_0_ to the infectious period *τ* (neglecting the latent period). This quantity is sensitive to the confinement measures and always lower with respect to time. We suppose that *r(t)* follows an exponential evolution, which we could justify by the fact that at early times it is much easier to reduce the infected-healthy contacts than at a late time. Therefore, we can write it as:

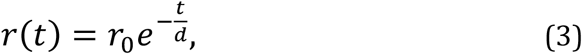

where *n* = *R*_0_*/τ* is the initial value calculated from the value of the reproduction number *R*_0_, which itself is estimated according to several considerations as 3.28-days in [12] or 6.47-days in [13]. Here, we will use the last one as a precaution to prevent bad decisions, thus *r*_0_ = 0.32. The parameter *d* is the number that characterizes the isolation strategy, which is -by definition-the number of days required to reduce the initial value *r*_0_ by 1/*e* ≃0.37. In our case, we must choose it as *d* = 341-days to get the simulation of detected cases compatible with the worldwide data of confirmed cases.

## 3 Results

### 3.1 Semi-closed infected population

Let us first start with a simulation to see what happens if we have a semi-closed infected population, where it is only possible to lose the infected without any gain. Therefore, let us fix the initial population to be *N*_0_ = 100 infected and extract every detected case following the *P_d_(t)* CDF. To do so, every time *t_n_= n*-days, we recalculate the numbers of infected and detected cases, where *t*_0_ = 0 is the moment of illness onset for the entire population. The number of infected *N(t)* at *t_n_* is then given by:

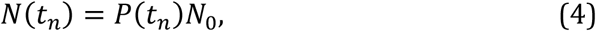

where we introduced a new distribution function *P(t*) written as follows:

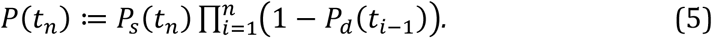

The number of detected cases *Ñ(t*) among the infected cases is given by:

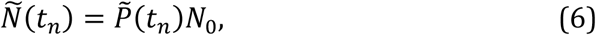

where 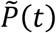 is a CDF function defined by:

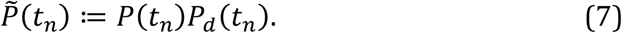

The behavior of these two numbers *N* and *Ñ* with respect to time -since the illness onset-is given in Fig: (4). It is noteworthy that the detected cases reach its maximum value near the incubation period and decrease afterward. The accumulation of detected cases is less than the initial number of infected *N*_0_ by a rate of 63% (the majority of infected are not detected).

**Figure 4:**
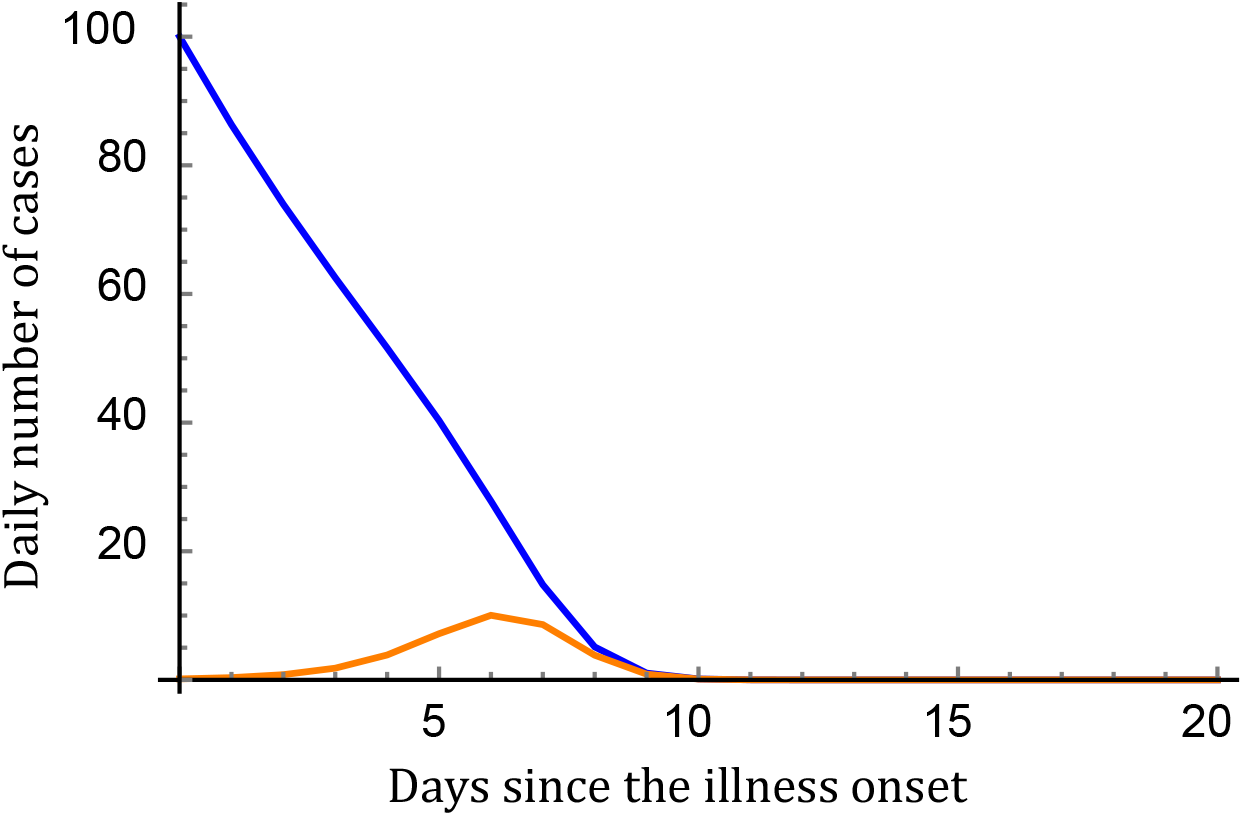
The graphs show the behavior of the number of infected cases *N(t)* (blue) and detected cases *Ñ(t)* (orange) in function of time

In the following section, we shall consider each “new cases” as a semi-closed population. Remark that after only 10-days there is no infected stay free, they are either isolated or recovered. For the last reason, we know that the period from the illness onset to death does not play a significant role in the detection mechanism.

### 3.2 Open population with an initial infected

Now, to apply the method to the worldwide status, the infected population must be considered as an open population where it is possible to gain and lose the infected. In essence, every day we must recalculate its remain cases as the following:

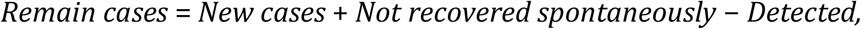

where we suppose that the new cases are given through the exponential growth [14]. Explicitly, if we suppose that the latent period [15] is zero so as to be able to multiply by *r(t)* since the first day of infection, we can then write it as:

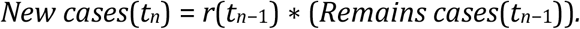

In general, we can write the true number of infected cases by a recurrence relation as:

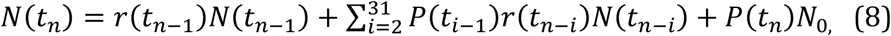

where *N(t_n-i_)* = 0 if *n <i*. The first term represents the new cases, the second term (formed by the sum of 30-terms) represents the cases that have not recovered spontaneously from the previously generated cases, and the last term represents the cases that have not recovered spontaneously from the initial number of infected (which of course is equal to zero if *t_n_*>30-days).

Likewise, the number of detected cases (confirmed) can be expressed as a recurrence relation as follows:

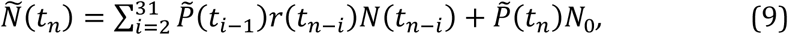

where the first terms under the summation symbol represent the detected cases from the previously generated cases and the second term represents the detected cases from the initial number of infected (which is equal to zero if *t_n_*>30-days).

Thus, using the above equations (8) and (9) by taking an initial number of infected cases as *N*_0_ = 270. Note that there were 27 cases at 15-Dec 2019 according to [16], and a detected rate claimed to be 10%. Thus, we can realize the simulation showing the evolution of the true infected cases and that of the detected one. The result is given by the two curves in Fig: (5)

**Figure 5:**
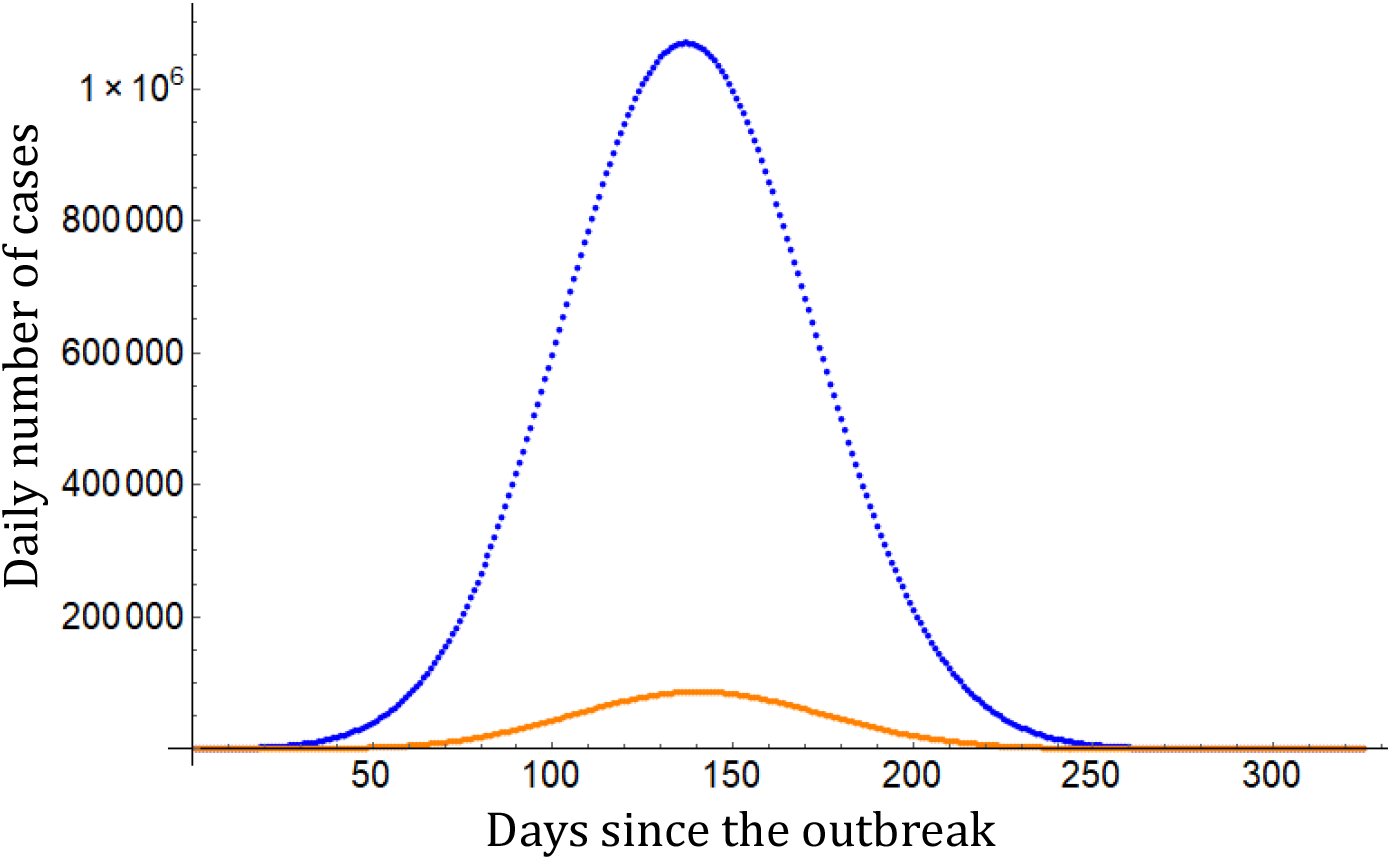
The evolution of the true infected cases *N(t*) (blue) and detected cases *Ñ(t*) (orange) in function of time

By looking at the two curves, we observe a great difference between them: the confirmed cases appear very smaller than the true infected cases. With a detection rate increasing across time (Fig: (6)). Beginning with an average of 5.3% in the first 50-days and ending by an average of 12% in the final 50-days before the disappearance of the pandemic. These results are well-matched with the results of the studies achieved in [17], [18]. In Fig: (6) we can see the daily evolution of the detection rate

**Figure 6:**
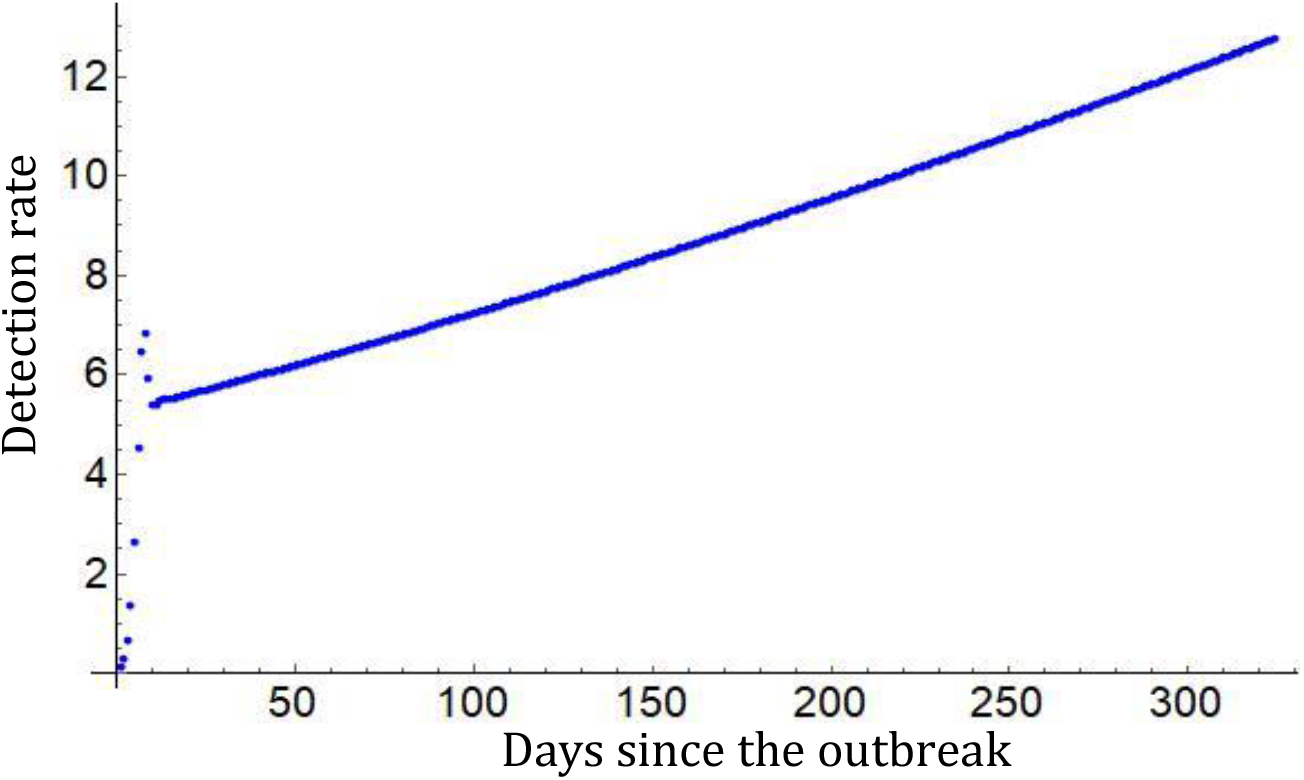
Detection rate evolution during the period of the pandemic

Finally, as we cited before, the curve of detected cases should be compatible with the confirmed cases as it is known from the available worldwide data [19]. This is what we do In Fig: (7), indeed, we compare the theoretical curve (dashed) with the real curve (continued) up to the present time.

**Figure 7:**
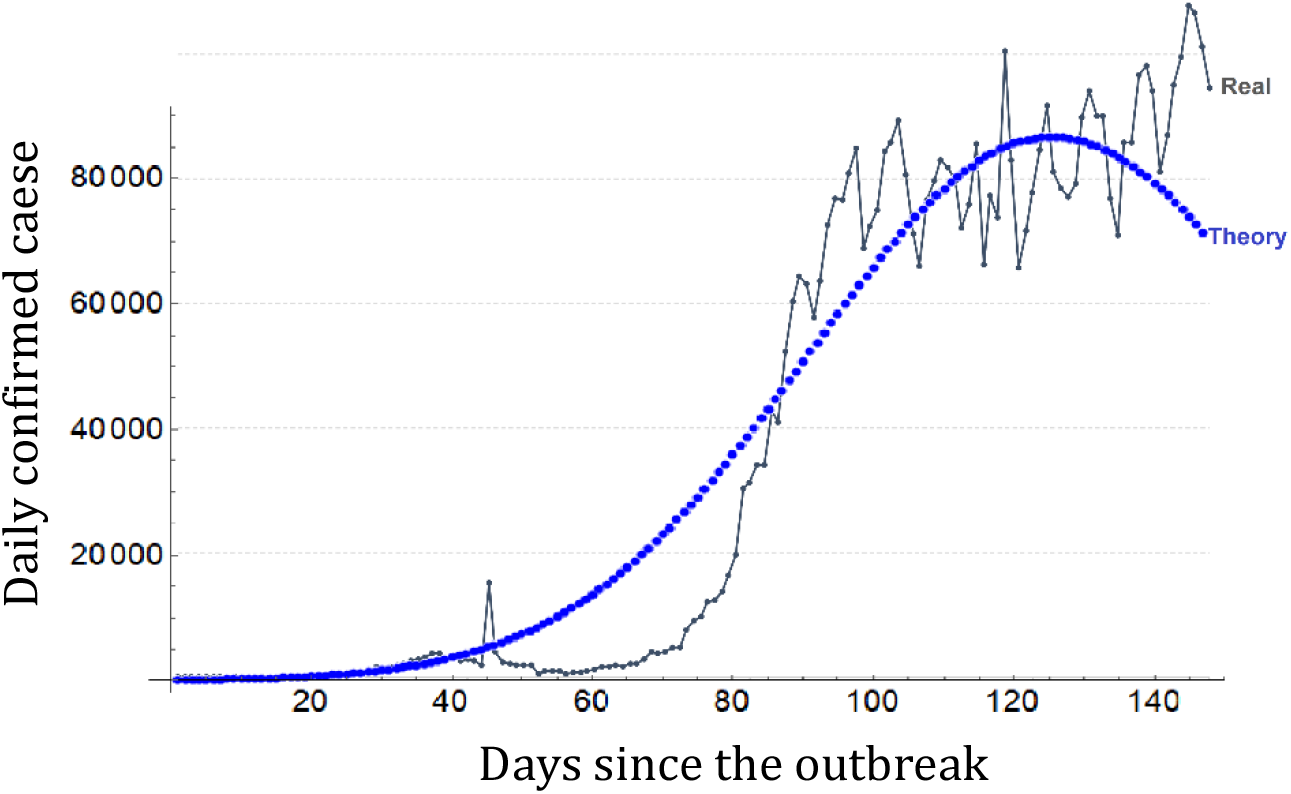
The graphs represent real (continued) and theoretical (dashed) curves of daily-confirmed cases.

Furthermore, we can deduce a numerical comparison between the simulated and the real accumulated -confirmed-cases ∑*Ñ(t*) which are 5.32M and 5.37M respectively (up to 25-May 2020 [19] In the other side, the true accumulated cases ∑*N(t)* give about 68.6M cases rather than 5.37M.

## 4 Conclusion

Across this work, we developed a new method to simulate the true evolution of the COVID-19 pandemic. This method consists of three concepts modeled by three analytic functions: the spontaneous outcome, the detected cases, and the well-known exponential growth. The results show in particular that the simulated curve of the true infected cases gives a large number in comparison with the confirmed one. The method is presented in a way that can be extended for use to other pandemics and/or to a given country. Furthermore, this simulation may help in predicting the impact of various measures taken to tackle the epidemic.

## Data Availability

The data that support the findings of this study are openly available in [our world in data], 19.

https://ourworldindata.org/

